# Optimizing blood pressure in acute intracerebral hemorrhage: protocol for a prospective individual participant data meta-analysis of the four INTERACT trials

**DOI:** 10.1101/2025.03.03.25323285

**Authors:** Xia Wang, Xinwen Ren, Qiang Li, Menglu Ouyang, Chen Chen, Candice Delcourt, Xiaoying Chen, Jiguang Wang, Thompson Robinson, Hisatomi Arima, Lu Ma, Xin Hu, Chao You, Gang Li, Jie Yang, Yapeng Lin, Laurent Billot, Paula Muñoz-Venturelli, Sheila Martins, Octavio Marques Pontes-Neto, Leibo Liu, John Chalmers, Lili Song, Craig S Anderson, the INTERACT Investigators

## Abstract

The Optimising Blood Pressure in acute intracerebral hemorrhage (ICH) study aims to conduct an IPDMA of pooling the four INTERACT trial to provide reliable information on the optimal blood pressure (BP) treatment target for patients with acute ICH. This protocol outlines the methods for the systematic review, research questions, and the approach to statistical analysis.

An intention-to-treat principle (ITT) will be applied in all analyses. Baseline characteristics will be summarised by treatment group. BP lowering treatment effect on the outcomes will be analysed by means of an ordinal logistic regression, and on the secondary and safety outcomes will be analysed by means of a binary logistic regression. Primary outcome is an ordinal distribution of modified Rankin Scale (mRS) scores at 90 days; secondary outcomes include change (absolute and relative, defined as an increase by >33%) in hematoma volume from the baseline and 24 hours CT scans and safety events within 90 days.

**Publication policy:** This policy is to be inclusive and follows that outlined for a prospective meta-analysis.^1^ This policy requires the identification of the lead investigator responsible for each randomized controlled trial (RCT) – known as a ‘Principle Investigator (PI)’– as well as up to four additional investigators from each RCT, members of the Trial Management Group or Trial Steering Committee for each RCT, and the methodologists/statisticians involved in the meta-analysis.

**Protocol:** Up to five investigators from each INTERACT trial may be included as co-authors of this protocol if they fulfil the ICMJE criteria of contributing authors, and they will be listed alphabetically by surname. The methodologists and statisticians involved in the design of INTERACT individual participant data meta-analysis will be co-authors. Each RCT may provide an unlimited number of non-author contributors, who are most likely to be members of each RCT’s Trial Management Group or Trial Steering Committee. The criteria for determining who is a contributing author or a non-author contributor follow ICMJE definitions: www.icmje.org/recommendations/browse/roles-and-responsibilities/defining-the-role-of-authors-and-contributors.html. All publications will name contributing authors as well as the attribution, “on behalf of the INTERACT collaborators.” Any report of the INTERACT IPDMA will be circulated to co-authors for comment and approval before submission for publication. All co-authors must have participated fully in manuscript preparation and editing.

## INTERACT trials

The INTERACT1^2^ and 2^3^ studies, and the third INTEnsive care bundle with blood pressure Reduction in Acute Cerebral haemorrhage Trial (INTERACT3),^4^ were international, multicentre, open, randomised controlled, blinded endpoint assessed, trials that involved adults with acute ICH within 6 h of the onset of symptoms and an elevated systolic BP (>150 mm Hg) who were allocated to receive intensive (target systolic BP <140 mm Hg within 1 h) or guideline-recommended (<180 mm Hg) treatment. In brief, INTERACT1 recruited 404 participants from hospitals in China, South Korea, and Australia in 2005-2007, and INTERACT2 recruited 2839 participants from hospitals in 21 countries in 2008-2012. The third INTEnsive care bundle with blood pressure Reduction in Acute Cerebral haemorrhage Trial (INTERACT3)^4^ used a multicentre, stepped wedge, cluster randomised, controlled trial design that incorporated early intensive BP lowering as part of a care bundle of protocols being implemented in 7036 patients at 121 hospitals in nine low-and-middle-income countries and one high-income country. Finally, INTERACT4^5^ randomly assigned patients with suspected acute stroke that caused a motor deficit and elevated systolic BP (≥150 mm Hg) within 2 h after the onset of symptoms, of whom 1029 participants had a haemorrhagic form of stroke, who were allocated in the ambulance to receive immediate BP lowering (target systolic 130 to 140 mm Hg within 30 min or usual BP management (usual-care upon arrival at hospital) at 51 hospitals in China.^6-8^

### Advantages of IPDMA to pool the four INTERACT trials

The four trials were all target-based international multicenter randomized controlled clinical trials, comparing intensive systolic blood pressure targets (<140 mmHg vs. <180 mmHg) to assess their impact on acute ICH outcomes. They implemented consistent treatment protocols using locally available antihypertensive agents, ensuring uniform intervention delivery across study sites. A standardized CT imaging protocol was adopted, with central expert review utilizing computerized multi-slice planimetric techniques to accurately measure bleeding volume. The trials also featured broad and simple inclusion criteria, allowing for robust generalizability while maintaining high-quality data collection through standardized assessments and rigorous quality control procedures. These methodological strengths reflect a harmonized study design and data collection approach, facilitating the generation of tailored insights for diverse populations and enhancing the power to detect small but clinically meaningful effects, ultimately contributing to more reliable and applicable clinical evidence in acute ICH.

### Data management

The data is stored on The George Institute (TGI) for Global Health’s Microsoft System Centre Data Protection Manager (DPM) server. TGI’s information technology (IT) infrastructure team manage a primary server, which takes a daily backup to a local disk pool which provides a daily snapshot recovery point with 42 days retention as well as a secondary DPM server in the Sydney Head Office which takes a daily backup of the data centre DPM’s most recent backup, thereby providing an offsite daily copy with 7 days’ retention and up to 90 days.

Data stored in TGI’s server will only be accessed from computers registered on the institute network using IT authentication service, or by using a virtual private network (VPN) connected to TGI when working from home at any time during the duration of the project lifecycle. TGI has in place system security SOP with VeriSign SSL digital certification and encrypted HTTPS connection (IT-SOP-105 v1.4). A data access and sharing policy has been published on the TGI website (https://sp.georgeinstitute.org/CORE/PoliciesAndSOPs/SOPs/DM-SOP-02%20External%20Data%20Sharing%20V1.0_signed.pdf). Data synthesis

We will check the range, completeness, and internal consistency of data items supplied by each trial. We will check that each RCT’s published aggregate results can be reproduced using the IPD supplied with reasonable accuracy, allowing for any data that are missing because participants opted out of data sharing.

### Aims

Aim 1: to determine the randomized BP lowering treatment effect on functional outcome at 90 days

Aim 2: to determine the randomized BP lowering treatment effect on hematoma growth at 24 hours post-randomization

Aim 3: to determine the randomized BP lowering treatment effect on safety outcomes

Aim 4: Pre-specified baseline patient characteristics (listed in table 1) will be explored to determine whether they modify the randomized BP lowering treatment effect

**Table 1.** Baseline characteristics of participants by randomised treatment.

| **Variable** | **Treatment group** |  | **All**<br>**N=xxx** |
| --- | --- | --- | --- |
|  | **Intensive BP lowering**<br>**(n=xxx)** | **Standard BP lowering**<br>**(n=xxx)** |  |
| Median time from onset to randomization, h |  |  |  |
| Mean age, y |  |  |  |
| Female |  |  |  |
| Region |  |  |  |
| Mean systolic BP, mm Hg |  |  |  |
| Mean diastolic BP, mm Hg |  |  |  |
| Median NIHSS score* |  |  |  |
| Median GCS score† |  |  |  |
| History of hypertension |  |  |  |
| Current use of antihypertensive drugs |  |  |  |
| Prior intracerebral haemorrhage |  |  |  |
| Prior ischaemic stroke |  |  |  |
| Diabetes mellitus |  |  |  |
| Use of warfarin anticoagulation |  |  |  |
| Use of aspirin/other antiplatelet agent |  |  |  |
| Haematoma location |  |  |  |
| Cortical |  |  |  |
| Deep‡ |  |  |  |
| Brainstem/cerebellar/primary ventricular |  |  |  |
| Median haematoma volume |  |  |  |
| Intraventricular extension of haemorrhage |  |  |  |
| Median intraventricular haematoma volume, ml |  |  |  |
| Median perihæmatomal oedema volume, ml |  |  |  |
| ICH score |  |  |  |
| 0 |  |  |  |
| 1 |  |  |  |

| Variable | Treatment group |  | All<br>N=xxx |
| --- | --- | --- | --- |
|  | Intensive BP lowering<br>(n=xxx) | Standard BP lowering<br>(n=xxx) |  |
| 2 |  |  |  |
| 3-5 |  |  |  |

Data are n (%), mean (SD) or median (IQR). BP=blood pressure, GCS=Glasgow coma scale, NIHSS=National Institutes of Health stroke scale
\*Scores range from 0 (normal) to 42 (coma with quadriplegia)
†Scores range from 15 (normal) to 3 (deep coma)
‡Location in the basal ganglia or thalamus

### Study outcomes

The following outcomes will be evaluated to ascertain the effects of early BP lowering treatment:

1. Primary outcome: an ordinal distribution of modified Rankin Scale (mRS) scores at 90 days.
2. Secondary outcomes: change (absolute and relative, defined as an increase by >33%) in hematoma volume from the baseline and 24 hours CT scans.
3. Safety outcomes: (i) early neurologic deterioration, defined as an increase from baseline of ≥4 points on scores of the National Institute of Health Stroke Scale (NIHSS) or a decrease from baseline of ≥2 points on the Glasgow Coma Scale (GCS), either within 24 hours; (ii) symptomatic hypotension requiring corrective therapy; (iii) death within 90-days; (iv) any serious adverse event (SAE); (v) any cardiac SAEs; and (vi) any renal SAEs.

## Data Availability

All data produced in the present study are available upon reasonable request to the authors

## Data analysis

The intention-to-treat principle (ITT) will be applied in all analyses. Baseline characteristics will be summarised by treatment group. BP lowering treatment effect on the primary outcome will be analysed by means of an ordinal logistic regression, and on the secondary and safety outcomes will be analysed by means of a binary logistic regression. Heterogeneity in the effects across the groups (listed in table 1) will be ascertained by adding interaction terms to the models.

To account for the heterogeneity of treatment effects across trials, a trial-by-treatment interaction term will be included in the models and dismissed if found to be non-significant. Sensitivity analysis, including additional covariate adjustments (baseline hematoma volume and National Institutes of Health Stroke Scale [<12 vs. ≥12)) will be conducted to assess the robustness of the results.

## Reporting

The reporting of this protocol complies with PRISMA-P10 and a recent recommendation.1 The results of this IPDMA will comply with the PRISMA-IPD statement,11 and mention identification of planned and ongoing RCTs, the IPDMA timeline, collaboration policies, and outcome harmonization processes.^1^

## References

1. Seidler AL, Hunter KE, Cheyne S, Ghersi D, Berlin JA, Askie L. A guide to prospective meta-analysis. Bmj 2019;367:l5342.

2. Anderson CS, Huang Y, Wang JG, et al. Intensive blood pressure reduction in acute cerebral haemorrhage trial (INTERACT): a randomised pilot trial. The Lancet Neurology 2008;7:391–399.

3. Anderson CS, Heeley E, Huang Y, et al. Rapid Blood-Pressure Lowering in Patients with Acute Intracerebral Hemorrhage. New England Journal of Medicine 2013;368:2355–2365.

4. Ma L, Hu X, Song L, et al. The third Intensive Care Bundle with Blood Pressure Reduction in Acute Cerebral Haemorrhage Trial (INTERACT3): an international, stepped wedge cluster randomised controlled trial. The Lancet 2023;402:27–40.

5. Li G, Lin Y, Yang J, et al. Intensive Ambulance-Delivered Blood-Pressure Reduction in Hyperacute Stroke. New England Journal of Medicine 2024;390:1862–1872.

6. Delcourt C, Huang Y, Wang J, et al. The Second (Main) Phase of an Open, Randomised, Multicentre Study to Investigate the Effectiveness of an Intensive Blood Pressure Reduction in Acute Cerebral Haemorrhage Trial (Interact2). International Journal of Stroke 2010;5:110–116.

7. Song L, Hu X, Ma L, et al. INTEnsive care bundle with blood pressure reduction in acute cerebral hemorrhage trial (INTERACT3): study protocol for a pragmatic stepped-wedge cluster-randomized controlled trial. Trials 2021;22:943.

8. Song L, Chen C, Chen X, et al. INTEnsive ambulance-delivered blood pressure Reduction in hyper-ACute stroke Trial (INTERACT4): study protocol for a randomized controlled trial. Trials 2021;22:885.

9. Hemphill JC, 3rd, Bonovich DC, Besmertis L, Manley GT, Johnston SC. The ICH score: a simple, reliable grading scale for intracerebral hemorrhage. Stroke 2001;32:891–897.

10. Moher D, Shamseer L, Clarke M, et al. Preferred reporting items for systematic review and meta-analysis protocols (PRISMA-P) 2015 statement. Syst Rev 2015;4:1.

11. Stewart LA, Clarke M, Rovers M, et al. Preferred Reporting Items for Systematic Review and Meta-Analyses of individual participant data: the PRISMA-IPD Statement. JAMA 2015;313:1657–1665.

